# Dynamic Changes in Prescription Opioids from 2006 to 2017 in Texas

**DOI:** 10.1101/19004879

**Authors:** Ebuwa T Ighodaro, Kenneth L McCall, Daniel Y Chung, Stephanie D Nichols, Brian J Piper

## Abstract

**Study Objective:** The US is experiencing an epidemic of opioid overdoses which may be at least partially due to an over-reliance on opioid analgesics in the treatment of chronic non-cancer pain and subsequent escalation to heroin or illicit fentanyl. As Texas was reported to be among the lowest in the US for opioid use and misuse, further examination of this state is warranted.

**Study Design:** This study was conducted to quantify prescription opioid use in Texas.

**Data Source:** Data was obtained from the publically available US Drug Enforcement Administration’s Automation of Reports and Consolidated Orders System (ARCOS) which monitors controlled substances transactions from manufacture to commercial distribution.

**Measurement and Main Results:** Data for 2006-2017 from Texas for ten prescription opioids including eight primarily used to relieve pain (codeine, fentanyl, hydrocodone, hydromorphone, meperidine, morphine, oxycodone, oxymorphone) and two (buprenorphine and methadone) for the treatment of an Opioid Use Disorder (OUD) were examined. The change in Morphine Mg Equivalent (MME) of all opioids (+23.3%) was only slightly greater than the state’s population gains (21.1%). Opioids used to treat an OUD showed pronounced gains (+90.8%) which were four-fold faster than population growth. Analysis of individual agents revealed pronounced elevations in codeine (+387.5%), hydromorphone (+106.7%), and oxycodone (+43.6%) and a reduction in meperidine (−80.3%) in 2017 relative to 2006. Methadone in 2017 accounted for a greater portion (39.5%) of the total MME than hydrocodone, oxycodone, morphine, hydromorphone, oxymorphone, and meperidine, combined. There were differences between urban and rural areas in the changes in hydrocodone and buprenorphine.

**Conclusions:** Collectively, these findings indicate that continued vigilance is needed in Texas to appropriately treat pain and an OUD while minimizing the potential for prescription opioid diversion and misuse. Texas may lead the US in a return to pre opioid crisis prescription levels.

## Introduction

Approximately one out of every twenty (4.6%) Texas survey respondents reported using opioid analgesics in 2009 nonmedically.^1^ Hydrocodone accounted for three-fifths of the calls involving opioids made to the Texas Poison Center Network between 2000 and 2010.^2^ Pill identification calls for hydrocodone in the Texas zip codes with large military bases increased from 2002 to 2009 by 463%.^3^ Opioid overdoses are generally lower than other states with similar reporting quality, however, overdoses involving synthetic opioids other than methadone increased by 28.6% from 2015 to 2016.^4^ Texas ranked 46^th^ in the US for per capita morphine mg equivalents (MME, 578) for ten prescription opioids and was approximately one-quarter the highest state (Rhode Island = 2,624).^5^ Texas had a similar ranking (45^th^) for buprenorphine and methadone for Opioid Use Disorder (OUD) treatment.^5^ Per capita calculations based on US Census data with the total population estimates should be interpreted with caution as undercounting, estimated at 239,500 in 2010 in Texas, may disproportionately impact minorities.^6^

There are several policy changes and demographic characteristics which may contribute to the rates of prescription opioid use and misuse. The Texas Prescription Monitoring Program, operated by the Texas State Board of Pharmacy, began collecting information about Schedule II prescription drugs in July, 1982 and Schedule III-V drugs in September, 2008.^2^ Texas implemented legislation to combat “pill mills” in September, 2010.^7^ The October, 2014 federal reclassification of hydrocodone from Schedule III to Schedule II by the Drug Enforcement Administration (DEA) was another policy change to combat the opioid crisis. A study that compared exposures from six-months before to six-months after the heightened regulation found that this restriction was followed by a decrease in hydrocodone exposures but also increases in codeine, oxycodone and tramadol reports to Texas Poison Centers.^8^ Texas had the highest percent (9.4%) of uninsured children (< 18) and uninsured adults (18-65, 19.0%) in 2015 in the US.^9^ The opioid epidemic has been described as iatrogenic.^5,10^ Economic disparities may, paradoxically, be protective at a population level against over-diagnosis or over-treatment with prescription opioids.^11^

Persons with non-white ethnicity also had lower drug poison deaths involving opioid analgesics.^12^ Perhaps the most important policy factor underlying prescription misuse is supply. The US Drug Enforcement Administration reduced opioid production quotas by ≥ 25% in 2017, particularly for hydrocodone.^13^ There have been concerns that rural patients are less likely to be able to access buprenorphine prescribers.^14^ Therefore, as Texas has been reported to be among the lowest in the US in opioid use^5^ and misuse^4,15^, there may be some insights from this populous and diverse state which could be beneficial for others. The goal of this pharmacoepidemiological investigation was to identify and quantify any changes in prescription opioids over the past decade in Texas.

## Methods

### Procedures

Drug manufacturers and distributors report controlled substance transactions to the DEA as required by the 1970 Controlled Substances Act. This information is made publically available by the Automated Reports and Consolidated Ordering System (ARCOS). ARCOS includes prescription data for Veterans Affairs patients, military personnel receiving care at non-Veterans Affairs pharmacies, Indian Health Services, dispensing practitioners (e.g. veterinarians) and Narcotic treatment programs (NTP) that may not be included in other databases. NTPs in Texas increased by 15.6% (77 to 89) from 2006 to 2017. Ten prescription opioids were selected based on prior research.^5,11^ Eight of these (hydrocodone, oxycodone, fentanyl, morphine, hydromorphone, oxymorphone, codeine and meperidine) are primarily used to relieve pain, and two (buprenorphine and methadone) are employed for an OUD. A high correspondence between ARCOS and a state prescription monitoring program for oxycodone (r = 0.99) was recently reported.^5^ Population data (28.3 million; 42.0% non-Hispanic white in 2017) was obtained from the US Census. Procedures were approved by the Institutional Review Board of the University of New England.

### Statistical Analysis

Three analyses were completed. First, the MME for each opioid was determined. Conversions were completed with the following multipliers: buprenorphine 10, codeine 0.15, fentanyl 75, hydrocodone 1, hydromorphone 4, meperidine 0.1, morphine 1, oxycodone 1.5, and oxymorphone 3.

Methadone from NTP had a conversion of 12 but 8 from other sources.^5,11^ Second, the weight in kg of each opioid was obtained for each year from 2006 to 2017. Percent change for each opioid, the two for an OUD, the nine primarily used for pain (including methadone from non-NTP sources), and all ten were calculated relative to 2006 and expressed as MME or kg of each agent. Third, heat maps of the percent change in the weight of hydrocodone or buprenorphine^14^ from 2012 to 2017 in each of the forty-nine three-digit zip codes reported by ARCOS were constructed with QGIS. Other figures were prepared with GraphPad Prism version 6.07.

## Results

The Texas population increased by 4.94 million between 2006 and 2017. Figure 1A shows that the MME of all opioids peaked in 2013 but only slightly (2.2%) exceeded the gains in population. Opioids for pain peaked in 2011 and subsequently returned to 2006 levels. In contrast, the agents employed to treat an OUD grew over four-fold faster than the population.

**Figure 1.**
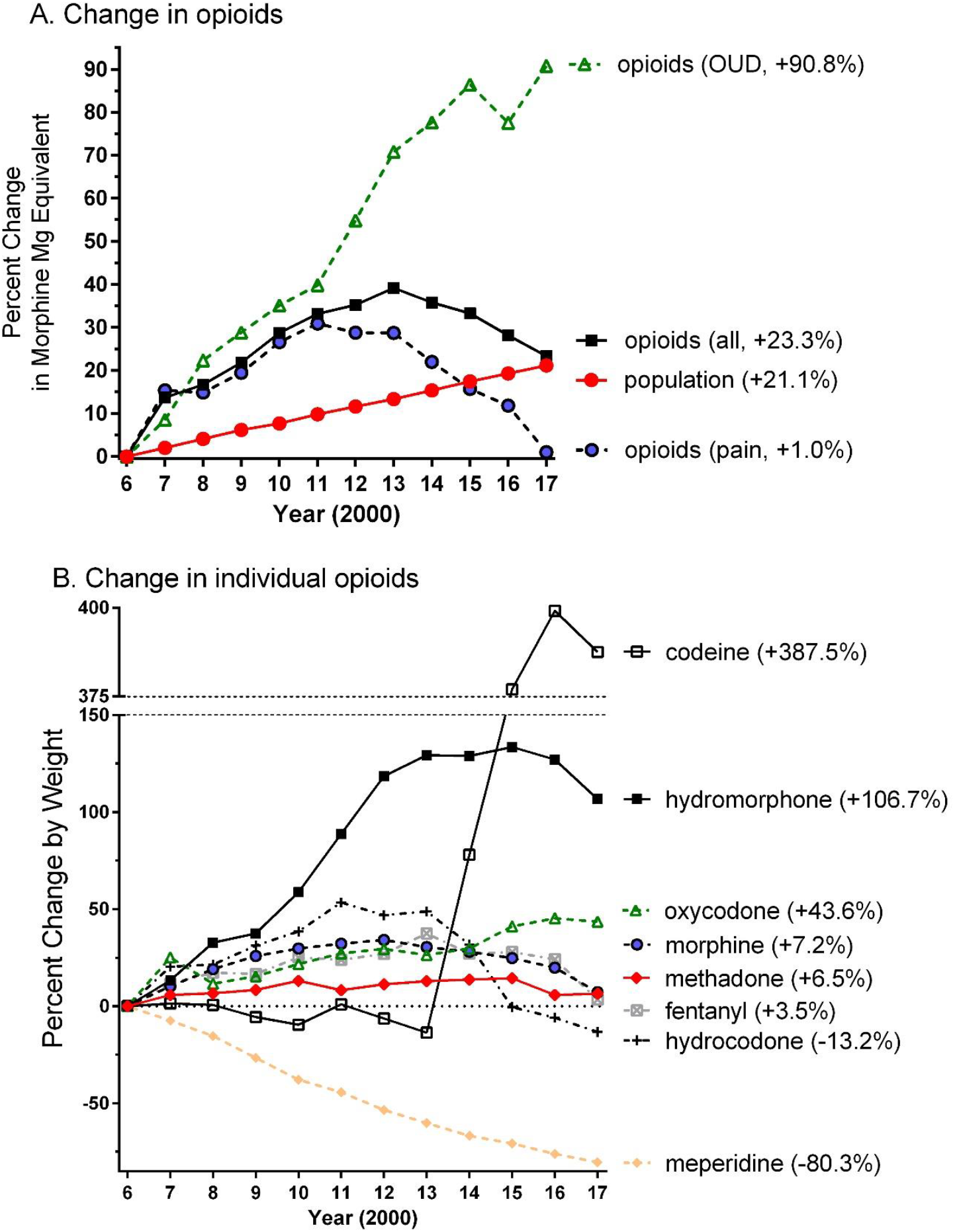
Percent change in weight, relative to 2006, of opioids used to treat pain (hydrocodone = 3,064.0, oxycodone = 922.2, codeine = 822.1, morphine = 765.8, methadone = 278.2, meperidine = 235.8, hydromorphone = 46.2, fentanyl = 22.5, oxymorphone = 3.3 kg), an opioid use disorder (OUD, methadone = 249.6, buprenorphine = 12.9 kg), or all (pain + OUD) by Morphine Mg Equivalent (A) or by weight (B) in Texas as reported to the Drug Enforcement Administration. Percent change versus 2006 is shown in parentheses.

Figure 1B depicts the changes in individual agents. Codeine showed a pronounced elevation from 2014 until 2017 and hydrocodone exhibited a reduction during this period. Hydromorphone doubled while the increases in oxycodone was double the population change. In contrast, meperidine showed a protracted decrease. Oxymorphone grew from 3.3 to 73.0 kg (+2,091.6%) in 2011, decreased slightly to 64.8 kg (+1,845.6%) in 2016, and to only 46.2 kg (+1,286.4%) in 2017. Methadone administered from non-NTPs (i.e. primarily pharmacies) was 278.2 kg in 2006, peaked at 293.5 kg in 2010, and declined to 153.2 kg in 2017. Methadone from NTPs grew by 64.0% from 249.6 kg in 2006 to 408.8 kg in 2017 (i.e. three-fold more quickly than population). Buprenorphine expanded, almost logarithmically, from 12.9 kg in 2006 to 105.3 kg in 2017 (not shown).

Figure 2 shows the percent of the total MME for each of ten-opioids in 2017. Methadone from NTPs was the top opioid and accounted for almost one-third of the total MME. Hydrocodone, oxycodone, and fentanyl combined were responsible for two-fifths (41.1%) of the MME.

**Figure 2.**
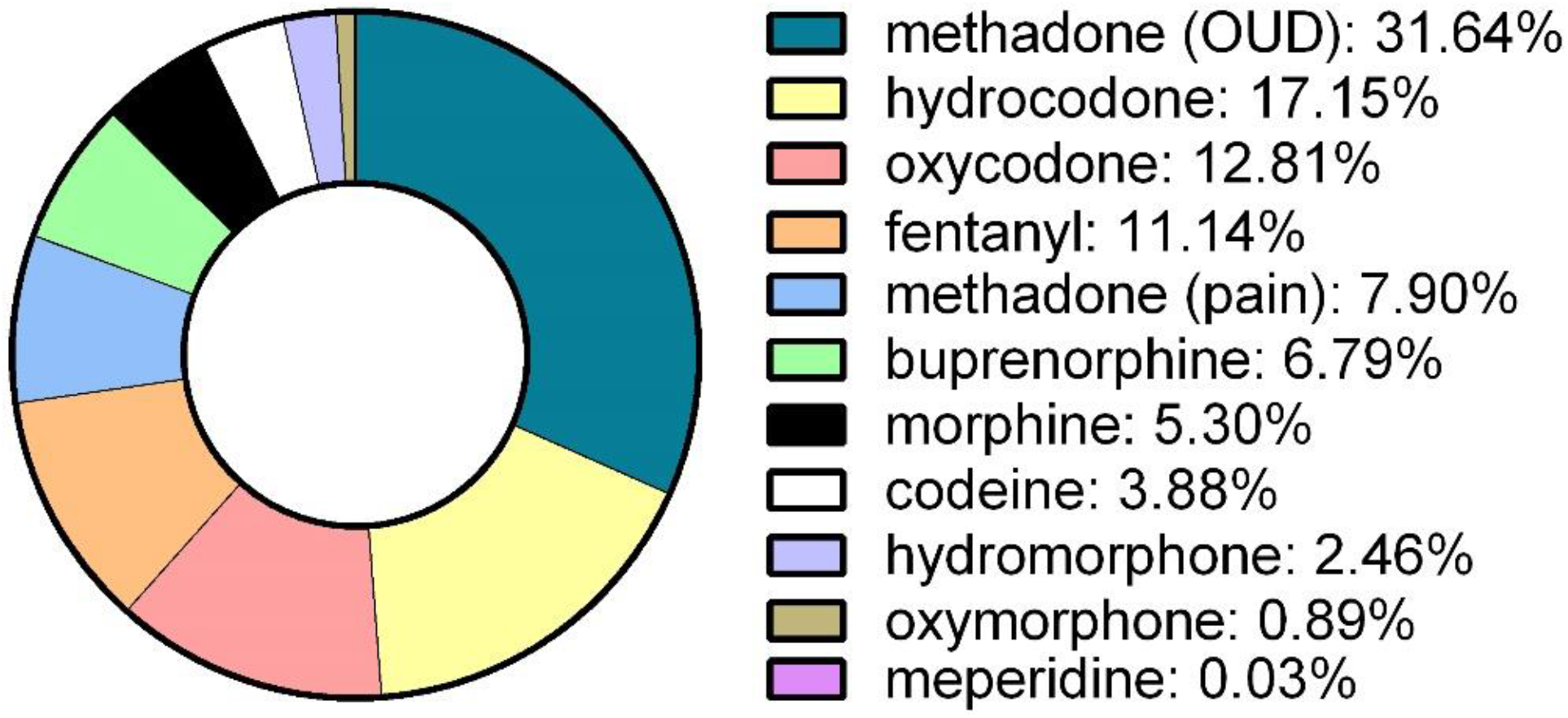
Morphine mg equivalents of ten opioids in Texas in 2017 as reported to the Drug Enforcement Administration’s Automation of Reports and Consolidated Orders System. Opioid Use Disorder: OUD.

Figure 3A shows the percent decrease in hydrocodone, by weight, in the last five years. With the exception of Amarillo and rural areas outside Lubbock and Austin, pronounced (≥ 32.5%) decreases were observed throughout the state. Figure 3B depicts that the percent increase in buprenorphine over the last half-decade was distributed across Texas.

**Figure 3.**
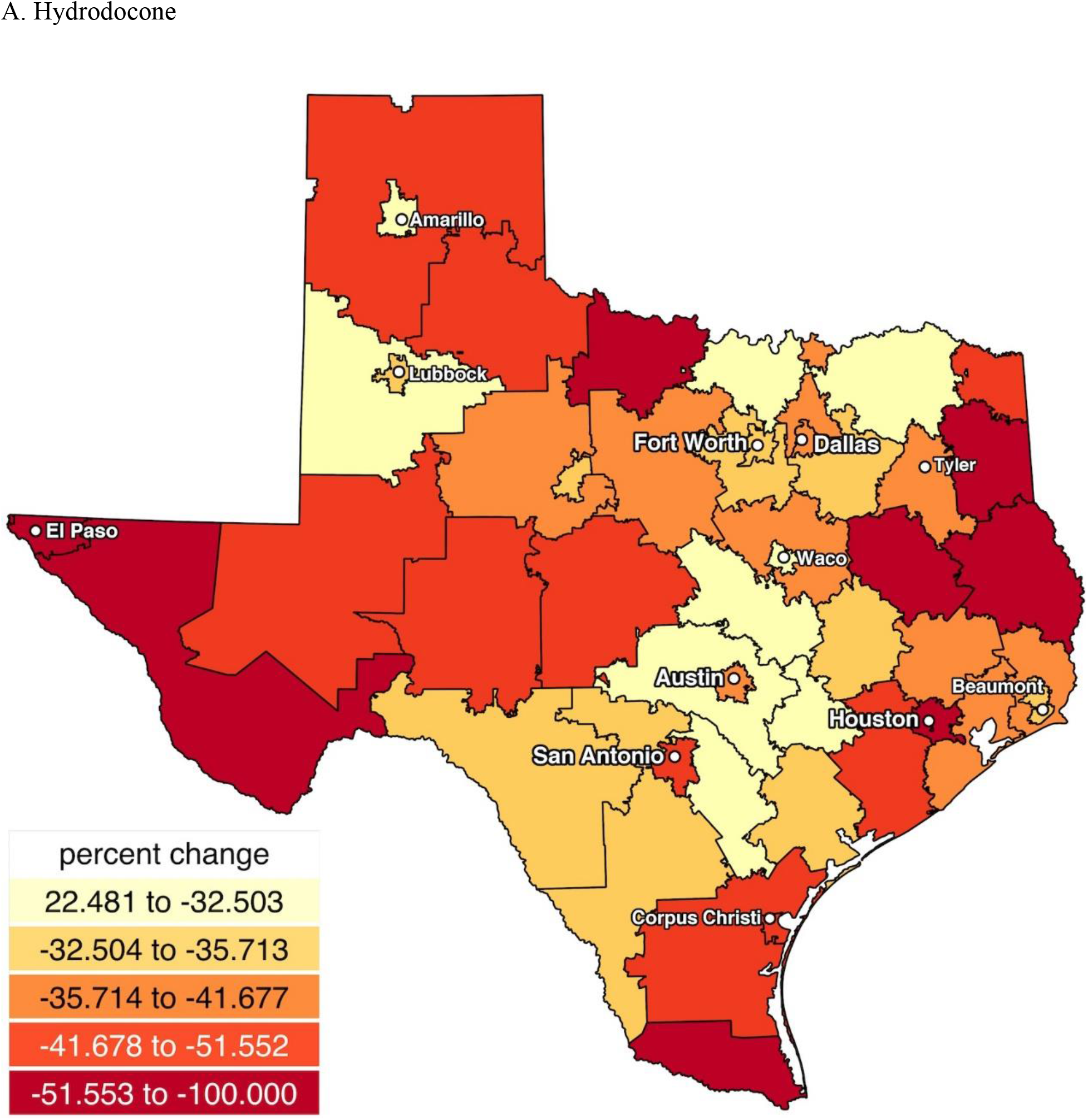
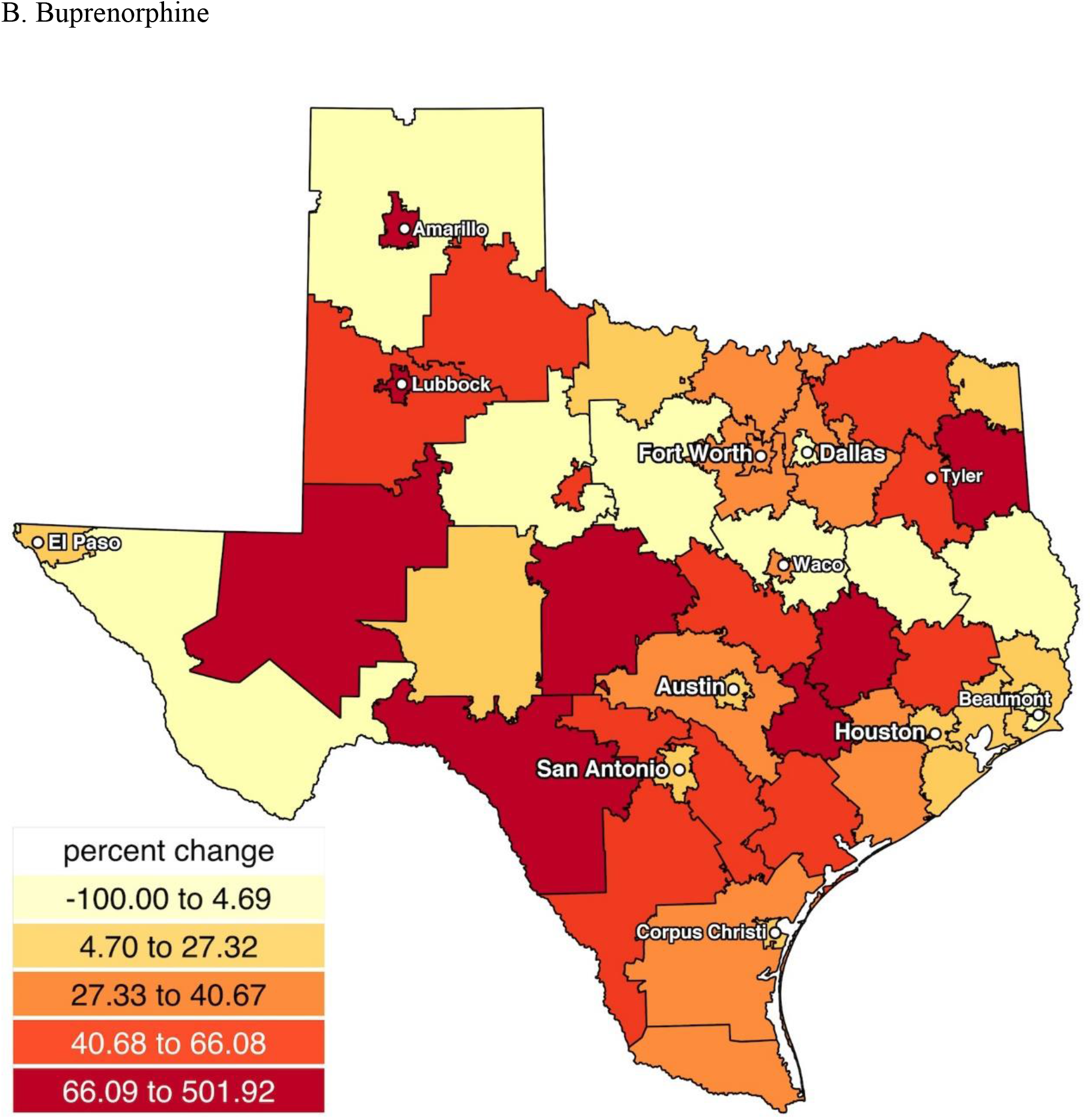
Heat map showing decreases in hydrocodone (A) or increases in buprenorphine (B) from 2012 to 2017 in Texas.

## Discussion

There were dynamic, pronounced and also agent specific changes in prescription opioids over the past decade in Texas. These results may be informative for other states or countries. The MME of all ten opioids peaked in 2013 and has been subsequently declining. Collectively, drugs used for the treatment of pain reached a maximum in 2011 and decreased each year thereafter to return to 2006 levels. Opioids for an OUD increased over ninety-percent. Nationwide, prior findings conducted either by the CDC^16^ or using ARCOS^5^ have found that prescription opioids for pain peaked around 2011 and have subsequently been declining.^17^ The rapid population growth in Texas may have delayed the temporal inflection point when more cautious opioid use for chronic non-cancer pain became evident.

Texas showed some parallels with the rest of the US when examining individual agents but also some differences. Meperidine showed a precipitous decline nationally^5^ and in Texas over the last decade. This is possibly due to increasing concerns relating to CNS excitability and seizure potential^18^ and the Libby Zion case.^19^ Concerns about its adverse reactions and potentially fatal drug interactions has prompted many to recommend restriction of its use or removal from the health-care system.^20^ Similarly, buprenorphine has rapidly increased in Texas and the US^5^ possibly due to concerns about methadone and NTPs^21^ and to increased access outside of NTPs, particularly in rural areas.^14^ Interestingly, the volume of codeine has been stable nationwide^5^ but showed a striking elevation in Texas. The rescheduling of hydrocodone had a clear impact (i.e. a 13.2% reduction occurring relative to a 21.1%% increase in population) and appears to be offset by a transition to other agents like codeine. These findings are congruent with those others.^17,22^ Percent increases and later reductions for oxymorphone should be interpreted within the context that the Opana ER formulation was only FDA approved for moderate to severe pain in 2006. The nationwide pattern is of a decrease in oxymorphone since 2011, likely resulting from the HIV cases in Indiana involving oxymorphone misuse.^23^ There are pronounced regional differences in use of the strong opioids hydrocodone and oxycodone. Hydrocodone was prescribed six times more commonly than oxycodone in Indiana.^10^ Oxycodone, by MMEs, was employed almost three-fold more than hydrocodone nationally.^5^

Texas has the unfortunate distinction of leading the country for most uninsured.^24^ The opioid epidemic has been described as iatrogenic.^5,10^ Prescription opioids were the first opioid among three-quarters of substance abuse treatment program admissions with heroin dependence.^25^ Possibly, economic factors could act as disincentives to limit chronic opioid use and decrease the likelihood of escalation to heroin or illicit fentanyl. Conversely, an absence of health insurance may contribute to an underutilization of evidence-based pharmacotherapies for an OUD like methadone^26^ or buprenorphine^27^ or to increased self-medication for pain, depression, anxiety, or sleep problems with non-prescribed opioids. Further national policy research is necessary to examine how variations in health insurance, including Medicaid expansion, may be associated with pharmacoepidemiological differences in the long-term treatment of pain or opioid misuse.

There are some strengths and limitations to this report and caveats of this data. ARCOS is a comprehensive data source in terms of including diverse patient groups often omitted from other studies and is publically available. For example, the Texas Prescription Monitoring Program is prohibited from reporting methadone or buprenorphine administered from NTPs. This is unfortunate because methadone from NTPs accounts for the most MMEs of any opioid in Texas and nationally.^5,11^ Persons with an OUD are currently denied the benefits of the Texas PMP due to 42 CFR Part 2. Unlike ARCOS, the Texas PMP may not consistently report on opioids from the Veterans Affairs, Indian Health Services, veterinarians, dentists, or Emergency Rooms. However, ARCOS does not report on all opioids that may be of interest (e.g. tramadol). The population information in Figure 1 is only as accurate as that reported by the US Census and is likely an under-estimate due to under-counting of undocumented persons. The illegal immigrant population nationwide was estimated to be 12.2 million in 2007 and decreased to 11.3 million in 2016.^28^ An accurate 2020 Census will continue to be important to inform public policy and health care planning. ARCOS tracks opioids from the point of manufacture to their point of sale or distribution at hospitals, practitioners, or retail pharmacies. State specific analyses may be underestimates if there were appreciable prescription opioids procured from out of state mail-order pharmacies. Although the increases in opioids for pain and an OUD exceeded the rapid rates of population elevations, ARCOS data is expressed in terms of overall drug weights. The body mass index of the population increased during the study period. One-quarter (26.3%) of Texas adults were obese in 2006 versus over one-third (33.0%) in 2017.^29^ Although opioids are not typically dosed on a body weight basis,^30^ obesity can contribute to lower back pain, osteoarthritis pain, and diabetic neuropathic pain. Other data sources will be needed to extend upon these results and identify changes in prescription opioid utilization among patients with specific indications.

In conclusion, state level analyses are important to better inform the contribution of both national and local policies to attenuate opioid misuse while preventing undertreatment for pain and insuring delivery of evidence based pharmacotherapies for an OUD. The Texas experience over the last decade with prescription opioids, particularly the dynamic changes with increases in buprenorphine and codeine but decreases in meperidine and hydrocodone, when coupled with population health indices,^2-4,7,8,14-16^ may offer some lessons for others to avoid or emulate.

## Data Availability

The raw data is available from the Drug Enforcement 
Administration at:
https://www.deadiversion.usdoj.gov/arcos/retail_drug_summary/index.html

https://www.deadiversion.usdoj.gov/arcos/retail_drug_summary/index.html

## Acknowledgements

BJP, DYC and SDN were supported by the Fahs-Beck Fund for Research and Experimentation. Software was provided by the Husson University School of Pharmacy and the National Institute of Environmental Health Sciences (T32 ES007060-31A1). Michael Sprintz, DO and Joseph Fraiman, MD provided feedback on earlier versions of this manuscript.

